# Effectiveness and cost-effectiveness of a culturally tailored Behavioural Activation intervention (DiaDeM) for treating Depression in Type 2 Diabetes: Protocol for a parallel arm, multi-country, randomised controlled trial in South Asia

**DOI:** 10.1101/2023.08.11.23294009

**Authors:** Faraz Siddiqui, Faiza Aslam, Naveed Ahmed, Saima Afaq, Asima Khan, Ada Keding, Simon Walker, Hannah Maria Jennings, Gerardo A Zavala, David Ekers, Edward Fottrell, Catherine Hewitt, Zia Ul Haq, Najma Siddiqi, DiaDeM Global Health Research Group

## Abstract

**Background:** The co-occurrence of depression among individuals with type 2 diabetes is a recognised global health problem and can lead to poorer health outcomes for both conditions. Behavioural activation is an evidence-based brief, low-cost psychological therapy which can be delivered by trained non-specialists, and is useful in treating depression, particularly in low-resource settings. The aim of this study is to test the effectiveness and cost-effectiveness of culturally adapted behavioural activation for depression in people with both depression and type 2 diabetes in two South Asian countries - Bangladesh and Pakistan.

**Methods:** A parallel arm, multi-country randomised controlled trial will be conducted in urban health care facilities providing diabetes services. We will recruit 604 adults in total, and randomise them using a 1:1 allocation ratio to receive culturally adapted behavioural activation (DiaDeM), or optimised usual care. DiaDeM comprises six sessions of behavioural activation with a trained non-mental health facilitator, conducted face-to-face and/or remotely. Optimised usual care includes information on depression, pharmacological and non-pharmacological treatment options for depression and details for accessing help locally. Participants in both arms will be followed up at 6- and 12-months post-randomisation. The primary outcome is the severity of depressive symptoms at 6 months, measured using the 9-item Patient Health Questionnaire (PHQ-9). Secondary outcomes include diabetes control, measured using glycosylated haemoglobin. An embedded process evaluation will evaluate the quality of intervention delivery and explore mechanisms of change and the contextual factors associated with the implementation and observed outcomes of DiaDeM. An economic evaluation will gauge DiaDeM’s cost-effectiveness and estimate the impact of diabetes and depression on economic outcomes.

**Conclusion:** There is an urgent need to address the rising burden of depression and chronic physical illnesses, such as type 2 diabetes. Interventions such as DiaDeM, which are culturally relevant and rely on a task-sharing approach, offer a potentially low-cost treatment within existing health services. If found to be effective and cost-effective, DiaDeM may be scaled up to address the mental health ‘treatment gap’ and improve mental and physical health outcomes for people with diabetes in South Asia.

**Trial registration:** ISRCTN40885204. Trial registered on 11th April 2023

## Background

Type 2 diabetes is a chronic non-communicable disease (NCD) associated with a disproportionately higher risk of having comorbid mental health conditions, particularly depression [1]. The two conditions have a bi-directional relationship - individuals with diabetes have double the risk of developing depression as compared to the general population [2–4], and having persistent depression also raises the risk of developing diabetes later in life [5]. Having depression alongside diabetes may impede diabetes self-care, causing a subsequent lack of control of overlapping risk factors such as diet and physical activity, and deterioration in the overall quality of life [2,3]. It also results in significantly higher healthcare use and expenditure as compared to diabetes alone [6]. These effects are likely to more severely impact populations in low-resource settings, particularly in low and middle-income countries (LMICs) where approximately 80% of individuals living with diabetes reside worldwide [7,8]. South Asia is a typical example of such low-resource, high-burden settings. Published literature from South Asian settings estimates the prevalence of diabetes to be as high as 26% [9], with at least a quarter of such individuals also having co-morbid depression [10,11].

Psychological therapies are increasingly being recognised for their relevance to the treatment of common mental disorders in people with diabetes, particularly because of their consideration of the wider psychosocial aspects of illness, and they have proven to be effective to reduce depressive symptoms in people with NCDs in LMICs [12]. Behavioural activation (BA) is a type of psychological treatment, recognised for its parsimony, portability across cultures and efficiency of training. It is a relatively simple, low-cost treatment, with established effectiveness for the treatment of depression in a range of settings and populations [13]. BA conceptualises depression in terms of the interaction between an individual and their environment; it helps people understand the link between mood and activities and encourages them to identify and engage in meaningful and rewarding activities as a means to improve depression [14,15]. BA treatment is widely accepted and is considered an economical alternative to, or adjunct to pharmacological treatment [16]. It can be effectively delivered by mental health staff, as well as trained non-mental health specialists [17,18]. Given these qualities, BA could be particularly relevant for LMIC populations and for priority groups such as those with co-occurring physical and mental Illnesses. It can overcome resource constraints in the provision of staged therapies and pharmacological interventions, and potentially contribute to reducing the burden of depression in people with diabetes. To date, only two clinical trials have evaluated the use of BA for the treatment of depression among individuals living with NCDs. Both were conducted in the United States, and only one demonstrated short and medium-term effectiveness in the remission of depression [19]. This evidence is limited, inconclusive and has little relevance for LMICs. Further high-quality studies are needed to evaluate the effectiveness and cost-effectiveness of BA for people with NCDs such as diabetes.

We have recently demonstrated the feasibility and acceptability of a culturally adapted BA intervention (the Diabetes and Depression Multimorbidity intervention - henceforth “DiaDeM”) to treat depression in people with diabetes in Bangladesh and Pakistan [14]. Building on this, we now aim to carry out a full-scale trial to test the effectiveness and cost-effectiveness of DiaDeM in these settings. Our objectives are to:

1. Evaluate the clinical effectiveness of the DiaDeM intervention in reducing the severity of depression at 6 months, compared with optimised usual care
2. Evaluate the clinical effectiveness of the DiaDeM intervention on secondary outcomes, compared with optimised usual care at 6 and 12 months
3. Evaluate the cost-effectiveness of the DiaDeM intervention over the trial period as well as the lifetime of affected individuals, considering the healthcare system and other broader perspectives
4. Estimate the causal impact of diabetes and depression on economic outcomes such as employment status, productivity loss, catastrophic health spending, household expenditure and household assets
5. Assess the fidelity and quality of intervention delivery, and identify the mechanisms of change and contextual factors associated with the implementation and outcomes of the DiaDeM intervention

## Methods

### Study design

The study is a multi-country, parallel arm, randomised controlled trial which will test the effectiveness and cost-effectiveness of the DiaDeM intervention compared to optimised usual care (control), and includes an embedded mixed-method process evaluation. The trial protocol (v1.0, dated 20/1/2023) was developed, and is reported in accordance with the Standard Protocol Items: Recommendations for Interventional Trials (SPIRIT) guidelines for clinical trial protocols (supplementary file 1) and the Template for Intervention Description and Replication (TIDieR) checklist (supplementary file 2) [20]. The DiaDeM trial was launched on 6th March 2023 and is currently in the recruitment phase. The activities related to trial recruitment, intervention delivery and follow-up assessments, and the overall flow of the trial are presented in Figures 1 and 2 respectively.

**Figure 1.**
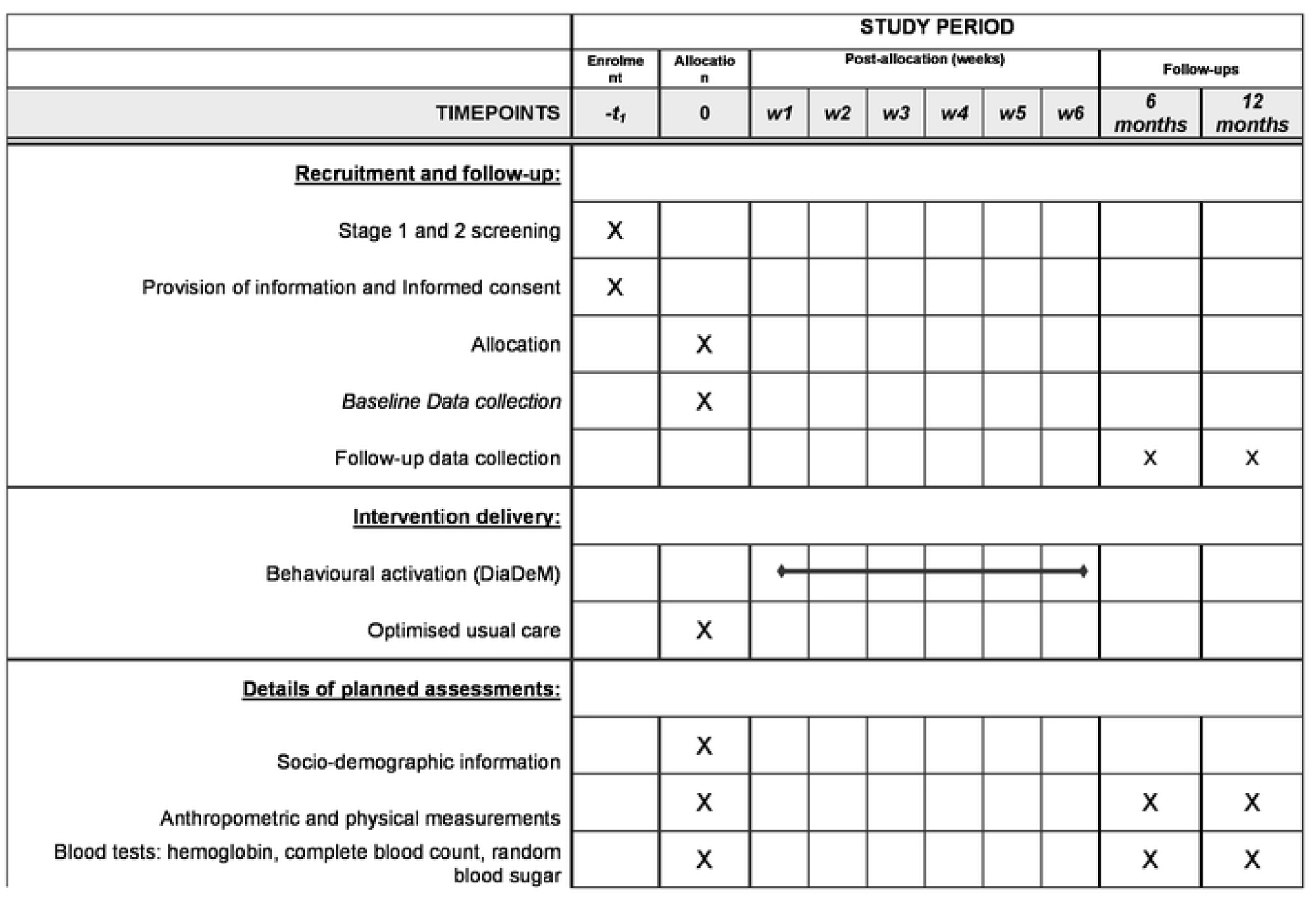
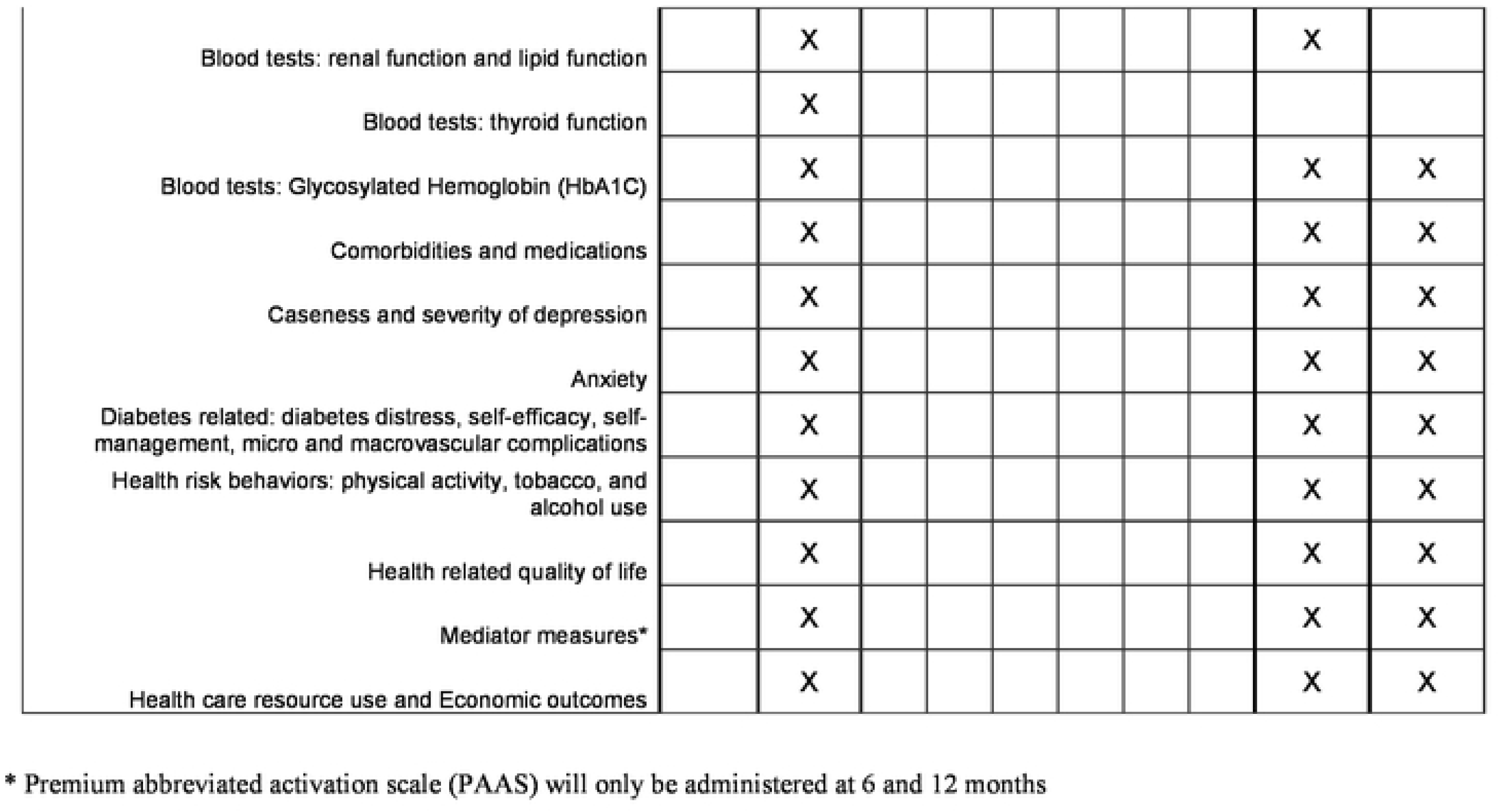
SPIRIT schedule of trial recruitment and follow-up, intervention delivery and assessments

**Figure 2.**
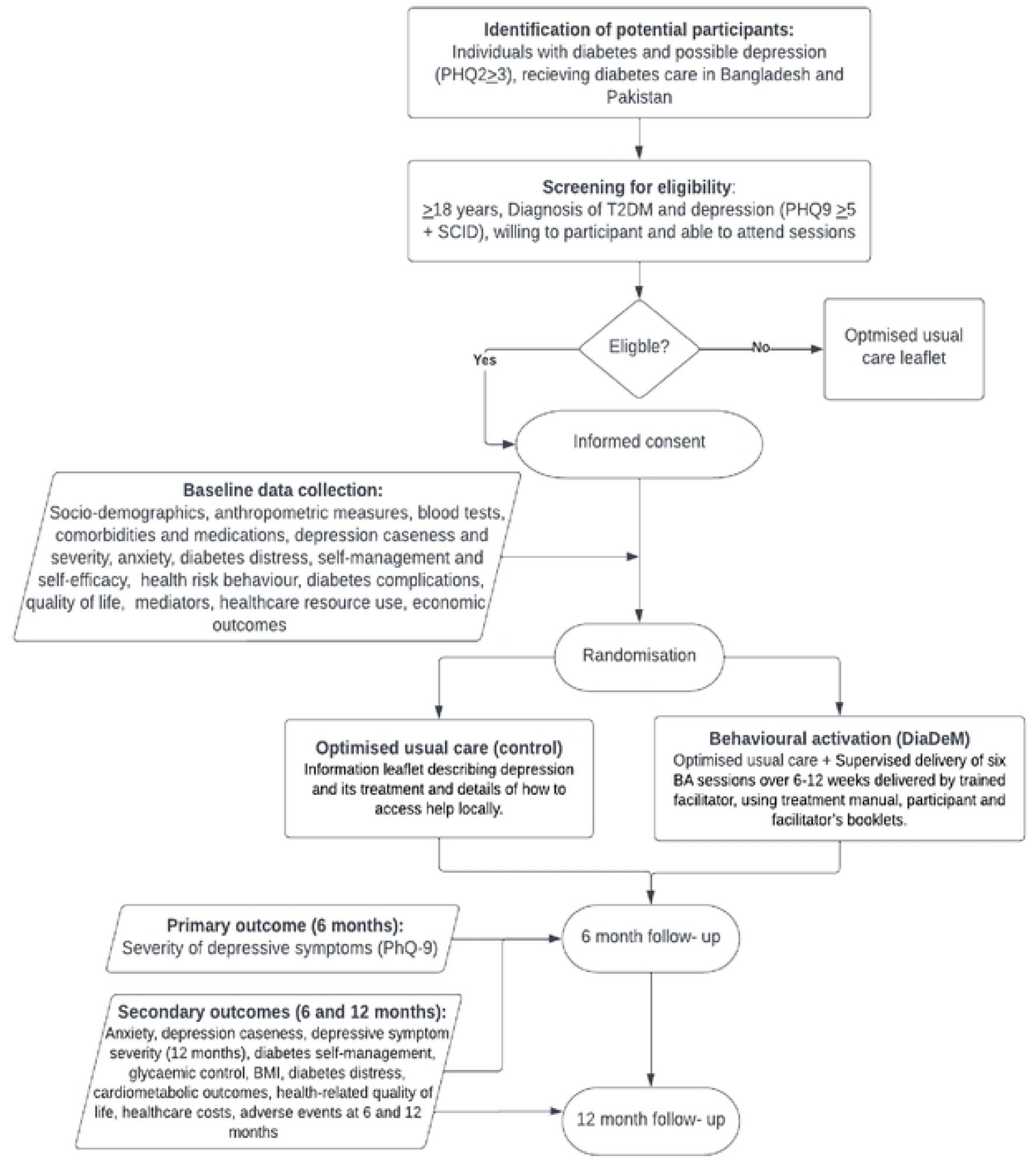
Flow diagram representing the recruitment, randomisation and follow up procedures of the DiaDeM trial

### Study settings and locations

The DiaDeM trial is being conducted in five urban health care facilities in South Asian settings that provide diabetes related services. Three of these sites are based in Pakistan (Karachi, Peshawar, and Rawalpindi) and two in Bangladesh (Dhaka, Sylhet). The chosen sites have well established diabetes services and available healthcare staff within these to deliver the DiaDeM intervention and have adequate research infrastructure. The sites are (in alphabetic order):

1. Baqai Institute of Diabetes and Endocrinology (BIDE), Karachi, Sindh, Pakistan.
2. Bangladesh Institute of Research and Rehabilitation in Diabetes, Endocrine and Metabolic Disorders (BIRDEM) General Hospital, Dhaka, Bangladesh.
3. Sugar Hospital, Phase V, Hayatabad, Peshawar, Pakistan
4. Sughra Diabetic Centre, Benazir Bhutto Hospital, Rawalpindi, Pakistan
5. Sylhet Diabetic Hospital, Sylhet, Bangladesh

### Eligibility criteria

This trial will be conducted in a sample of individuals who have a confirmed diagnosis of type 2 diabetes and depression and are seeking outpatient diabetes care in the above-mentioned study sites in Bangladesh and Pakistan. Eligible individuals will be identified based on the following criteria:

- Age >18 years old at the time of screening/recruitment
- Confirmed physician diagnosis of type 2 diabetes based on standardised diagnostic criteria (clinical presentation and HbA1C levels) and currently receiving treatment at the diabetes centre
- Scoring ≥3 on the Patient Health Questionnaire-2 (PHQ-2)[21] depression screening tool, and subsequently scoring ≥ 5 on the Patient Health Questionnaire-9 (PHQ-9)[22] with depression confirmed using the current major depressive disorder section of module A “Mood Episodes and Disorders” of the Research Version of the Structured Clinical Interview for the DSM-V (SCID-V-RV)[23]
- Willingness to attend BA sessions in person or remotely

Individuals in any of the following categories will not be eligible for trial participation.

- Currently receiving psychotherapy for depression at the time of screening/recruitment
- Unable to provide consent due to cognitive impairment or severity of psychological or physical illness

### Screening and identification of eligible participants

Eligible participants will be identified using a two-stage process. In the first stage, patients attending outpatient clinics at each study site will be screened for depressive symptoms by clinical staff, using the two-item Patient Health Questionnaire (PHQ-2). Positively screened individuals will be provided information about the importance of depression screening (leaflet 1) (supplementary file 3) and where to access help; those who agree to further screening will be taken to phase 2 following written consent for the screening of depression (Supplementary file 4). Each individual will be assigned a unique screening ID at this stage and will be assessed further by a research assistant (RA) for depression using the Urdu or Bangla versions of the nine-item PhQ-9 followed by the Current Major Depressive Disorder section of module A of the SCID-V.

The RAs will check, confirm and record the eligibility of the identified individuals for trial participation. Those who score positive on both PHQ 9 (score ≥5) and SCID-V and fulfil the remaining eligibility criteria described above will be taken forward for full consent for trial participation.

### Informed consent and recruitment

RAs will provide eligible individuals with a local language (Urdu or Bangla) version of the participant information sheet (Supplementary file 5), which provides details about the trial, its methods and assessment procedures, as well as any possible advantages and risks associated with participation. The opportunity to read the information sheet will be given to eligible participants; those who are unable to read will have the information sheet read out to them. Participants will be made aware of their rights, including the right to withdraw from the study at any time and the procedure to follow in such a situation.

After the information sheet has been read, the RAs will provide a study consent form (Supplementary file 6) to the participants to read and confirm their participation. Written consent will be recorded. Following consent, the recruited participant will be taken forward to baseline data collection and then randomised. The participant’s consent status and contact information will be recorded in a participant log and an electronic database.

### Randomisation and blinding

After completion of the baseline Case Report Form (CRF), each recruited participant will be randomly allocated to either the treatment or control arm. Randomisation will use a 1:1 allocation ratio and will follow a computer-generated randomisation sequence that will be stratified by country with randomly permuted blocks of varying block sizes, generated using Stata version 17 or later (StataCorp, 4905 Lakeway Drive, College Station, Texas, 77845 USA). A request for randomisation will be made to the independent statistician based at the University of York. This will be done via email by the site RA or by the country data manager in case of internet disruption at the study site. An automated email will be sent back to the RA and data manager containing the trial ID and allocation. The trial ID will be recorded on the completed baseline CRF and the recruitment log. The trial manager and the RAs collecting outcome data at the follow-up stage will be blinded to participant allocation status.

### Details of Intervention and Control

#### Behavioural activation (DIADEM)

The DiaDeM BA intervention is a culturally adapted, brief, psychological therapy for depression which focuses on behaviour change to enhance mental and physical well-being for people living with depression and diabetes multimorbidity. The intervention was adapted from the Multimorbidity in older adults intervention (MODS) [24] programme using the intervention mapping framework for the development, implementation and evaluation of health promotion interventions [25]. This framework provides a structured approach for the development and evaluation of health interventions. Additionally, the Bernal [26] and Escoffery [27] frameworks were employed to specifically address the cultural components of the adaptation process. These frameworks facilitated a comprehensive and systematic adaptation of DiaDeM, ensuring that it was culturally relevant and appropriate for the South Asian population. As part of the adaptation process, we developed a logic model based on the desired outcome “to improve depression in people with diabetes in Bangladesh and Pakistan”. To understand the “mechanisms of action” through which the intervention affects behaviour, we identified behavioural change techniques and the mechanisms of action they target. Full details of the adaptation process are described in detail elsewhere [28].

DiaDeM will be delivered by non-mental health specialist staff (diabetes staff working as nutritionists, health educators, nurses, or paramedics etc). The non-mental health specialist BA facilitators (referred to henceforth as ‘BA facilitators’) will be guided and supervised by mental health specialists in Bangladesh and Pakistan who will be referred to as ‘BA supervisors.

DiaDeM comprises six weekly sessions (each lasting 30-45 minutes) delivered by a BA facilitator using the BA facilitator’s manual. Session one will introduce the concept of the relationship between mood and behaviour and between purposefully scheduling and engaging in rewarding activities in improving symptoms of depression and self-management of diabetes. Sessions two to five are skills-based for learning and practising skills that support the participant in re-engaging in personally chosen, rewarding behaviours. Each of these sessions incorporates a recap of learning from the previous session and a review of planned between-session work. Participants are supported by the BA facilitator to increase both the number of rewarding behaviours engaged in week-by-week and to gradually introduce behaviours previously considered too difficult to manage as their symptoms of depression improve. Session six consolidates prior learning and promotes maintenance behaviours by developing a personalised “staying well” plan. The participants will also be given a ‘participant booklet’, a take-home resource that incorporates key messages from the intervention sessions as well as space in which to record information about between-session behaviour change activities to practice, chosen by each participant with the support of the BA facilitator. DiaDeM sessions will be delivered individually in private treatment rooms at each study site. If the participant is unable to travel to the treatment centre, they will be offered a video call or telephone treatment session by the BA facilitator as an alternative.

BA facilitators and the supervisors will receive prior training in using the DiaDeM manual and participant booklet to support DiaDeM treatment sessions and in supporting participants in choosing between-session behaviour change activities. All the BA facilitators and supervisors will attend an online, introductory course on Behavioural Activation for Depression developed by the University of York (https://www.impactsouthasia.com/behavioural-activation-learning/) as a requisite for attending the BA training. A three-day hands-on face-to-face BA training will be conducted in Bangladesh and Pakistan by BA experts and therapists. Training will be conducted for both facilitators and supervisors jointly, using PowerPoint presentations, individual and group activities, and role-play. The supervisors will receive additional training on oversight, after which they will perform the competency assessments of BA facilitators. During the intervention delivery phase of the trial, the supervisors will oversee and guide the BA facilitators and will conduct refresher training, if required. The supervisors will also record supervisory notes and competency assessments in “supervisor’s logs”; BA facilitators will record details of the BA delivery sessions for each participant in the facilitator’s logs.

#### Optimised usual care

Optimised usual care comprises information leaflets on depression, pharmacological and non-pharmacological treatment options for depression and the procedures for accessing local help, provided by the study team. The leaflets were developed by the project team in consultation with mental health experts (Supplementary file 8). The intervention group will also receive optimised usual care.

### Sample size

The sample size was calculated in Stata (Version 17) and is based on sufficient power for the analysis of data in each country, although data will be combined for the primary analysis. A meta-analysis of Behavioural Activation for depression reported a standard effect size of -0.74 (95% CI of -0.91 to -0.56) [29]. The lower bound value of 0.5 is equivalent to a difference of approximately 2 PHQ-9 points (SD of 4). The feasible number of intervention facilitators differs in each country setting, resulting in different potential clustering effects to account for.

In Pakistan, in order to detect the above difference with 90% power and 5% alpha, accounting for clustering by therapist (ICC of 0.04, average of 10 intervention participants per therapist) and assuming 20% attrition, a total of 294 participants is required to be recruited (147 participants in each arm). A minimum of 15 intervention facilitators is required. In Bangladesh, in order to detect the same difference, assuming an average of 12 intervention participants per therapist (minimum of 13 intervention facilitators), a total of 310 participants are required to be recruited (155 in each arm).

This sample size estimate is conservative, applying the clustering inflation factor to both study arms to maintain the randomised allocation ratio of 1:1. The achieved power for the recruitment target in each country based on clustering in the intervention arm only exceeds 97% (clsampsi command in Stata). The combined total sample size for both participating countries is 604. Assuming a conservative estimate of depression prevalence of 10% and a 60% consent rate, approximately 10,070 patients will need to be screened.

### Primary and secondary trial outcomes

The primary trial outcome will be the severity of depressive symptoms, assessed using the PHQ-9 instrument at 6 months post-randomisation. This is a nine-item questionnaire scored from a minimum of 0 to a maximum of 27 [22,30]. Secondary trial outcomes are depression (caseness), anxiety, diabetes self-management, glycaemic control, BMI, diabetes distress, cardiometabolic outcomes, health-related quality of life, healthcare costs, adverse events at 6 and 12 months; severity of depressive symptoms at 12 months. Details of how and when these outcomes will be measured are provided in the text below.

### Baseline and follow-up data collection

Baseline participant data will be obtained before randomisation at the time of recruitment. Follow-up participant data will be collected 6 months and 12 months post-randomisation. RAs will use electronic CRFs to collect information either face-to-face, or remotely (in case of COVID-19 restrictions, or if not feasible to travel) at all time points. To reduce the risk of unbliding the treatment allocation, the RAs will be switched at follow-up time points. The details of each planned assessment are provided below, and the time-points at which these will be collected are presented in Figure 1.

### Socio-demographic information

The WHO Stepwise approach to surveillance (STEPs) instrument, Version 3.2 [31] will be used to collect information about participants’ age, sex, education and marital status. Translated versions of this instrument have been used and validated in different countries of South Asia including Bangladesh [32] and Pakistan [33].

### Anthropometric and physical body measurements

The following physical body measurements will be taken by trained RAs following the protocols elaborated in the WHO manual [34].

- Weight (in kilograms) will be measured using a portable digital weighing scale will be used at all sites. The weighing scale will be placed on a flat surface - two repeated readings will be collected and recorded to two decimal places. Participants will be instructed to wear light clothing and to remove footwear and socks at the time of weighing.
- Height (in centimetres) will be measured using a portable stadiometer, without footwear and headgear. Two consecutive readings will be recorded to 0.1 cm.
- Body Mass Index will be calculated as weight in kilograms (kgs) divided by height in meters squared (kg/m^2^). and recorded in the electronic tablet.
- Waist circumference (in centimeters) will be measured at the midpoint between the lower margin of the last palpable rib and the top of the iliac crest (hip bone) with a relaxed arms position and fully exhaling process at the end of expiration. Two repeated readings will be recorded with a precision of 0.1 cm.
- Hip circumference (in centimeters) measurement will be recorded using a flexible anthropometric tape by wrapping the tape around the widest part of the buttocks, and taking the parallel to the floor. Two repeated readings will be recorded in centimetres with a precision of 0.1 cm.
- The waist-hip ratio will be calculated by dividing the waist circumference by the hip circumference, and recorded on the electronic tablet.
- Blood pressure and heart rate will be measured with the help of an automated blood pressure measuring instrument. Three consecutive readings will be recorded (each with a 3-minute gap) from the same wrist/elbow and using the same device. The participant will be comfortably seated at the time of conducting the measurement. After completing the whole process participants will be informed about the readings. The measurement of blood pressure will be recorded from the left wrist/arm but in the case of a stroke on that side/limb then it will be measured from the right side.

### Blood tests

Blood samples of all the study participants will be drawn at each study site for the biochemical assessment after completion of all the measures using standardised protocols. The blood samples in Pakistan will be transported and investigated at the standardised pathological clinical laboratory of Baqai Institute of Diabetes and Endocrinology, Karachi. In Bangladesh, samples collected for the glycated hemoglobin (HbA1C) will be transported for analysis at the BIRDEM General Hospital, Dhaka. Glycated haemoglobin (HbA1c) will be assessed at baseline, 6 and 12 months follow-ups. Hemoglobin level (Hb), Complete Blood Count, Random Blood Glucose (RBG) test, renal function test including serum creatinine and estimated Glomerular Filtration Rate (eGFR), lipid function test including Total Cholesterol (TC), High-Density Lipoprotein Cholesterol (HDLC) and Low-Density Lipoprotein Cholesterol (LDLC) will be assessed at baseline and 6 months. Thyroid-Stimulating Hormone (TSH) test will be performed only at baseline.

### Comorbidities and medications

Data on comorbidities and medications will be collected using participant self-report, through a set of questions adapted from the STEPs module for NCDs [35]. Participants will be asked for a medically diagnosed history of raised blood pressure, heart disease, hypercholesterolemia, stroke, lung diseases, the treatment offered by their health workers and communicable/ infectious diseases (e.g. Tuberculosis, Malaria, HIV, Hepatitis B, C, Dengue, COVID-19, Chikungunya).

### Caseness and severity of depression

The Patient Health Questionnaire-9 (PHQ-9) is a nine-item questionnaire and scored from a minimum of 0 to a maximum of 27, with a cut-off of 0–4 represents no depressive symptoms, 5–9 mild depressive symptoms, 10–14 moderate depressive symptoms, 15–19 moderately-severe depressive symptoms, and 20–27 severe depressive symptoms [22,30].

### Anxiety

The severity of generalised anxiety disorder will be assessed with the use of a generalised anxiety disorder (GAD-7) [36,37] scale. It is a 7-item scale with scores ranging from 0 to 21, where the scores of 5, 10, and 15 will be taken as cut off points for mild, moderate, and severe anxiety [36].

### Diabetes-related distress

The Problem Area in Diabetes Scale-5 (PAID-5) will be used to collect information on diabetes-related emotional distress [38,39]. This scale will assess common issues and feelings linked with complications of diabetes. The 5-item scale scores range from 0 (not a problem) to 4 (serious problem). The total sum of scores of the PAID scale determines the level of diabetes-related distress and a cut-off of ≥8 indicates diabetes-related distress [38].

### Diabetes-related self-efficacy and self-management

Three measures will be used: the Diabetes Empowerment Scale Short Form (DES-SF)[40], Perceived Diabetes Self-Management Scale (PDSMS) [41] and Summary of Self-Care Diabetes Activities scale (SDSCA) [42]. The DES-SF is an 8-item scale to assess general diabetes-related psychosocial self-efficacy. The 8-item PDSMS measures confidence in self-management with individual scores ranging from 1 (strongly disagree) to 5 (strongly agree) with 4 items being reversed scores. The total score of the PDSMS scale ranges from 8 to 40, with higher scores indicating higher confidence in self-management. The 11-itemSDSCA measures diabetes self-management activities related to general diet, specific diet, exercise, blood glucose testing, foot care and smoking).

### Health risk behaviours

Information on physical activity will be collected using the International Physical Activity Questionnaire (IPAQ), a nine-item scale (short version) [43]. The scale measures activities and their intensity over the past seven days i.e. 1) vigorous-intensity activity such as aerobics, 2) moderate-intensity activity e.g. leisure cycling, 3) walking and 4) sitting. Current use of smoked or smokeless tobacco in the past 30 days will be assessed through questions adapted from GATS questionnaire and the STEPs expanded modules on tobacco and smokeless tobacco use [31]. Alcohol use: Questions related to alcohol use, adapted from the STEPs module [31] will also be applied to measure alcohol consumption.

### Diabetes-related microvascular and macrovascular complications

Current or previous episodes of macrovascular complications (coronary artery disease, stroke and peripheral arterial disease) along with microvascular complications of diabetes (diabetic nephropathy, neuropathy and retinopathy) will be assessed based on health care staff reporting or by participant’s self-report.

### Health-related quality of life

Euroqol’s instrument EQ-5D-5L will be administered to measure health-related quality of life (HRQoL) [44]. EQ-5D is a standardised measure of health status developed by the EuroQol Group. It provides a simple, generic measure of health for clinical and economic appraisal, where health is characterised by five dimensions; mobility, self-care, ability to undertake usual activities, pain/discomfort, and anxiety/ depression [45]. participants’ subjective evaluation of their health state based on a visual analogue scale (EQ-5D-VAS) is also included in the scale, ranging from 0 (representing the worst imaginable health state) to the maximum attainable value of 100 (indicating the best imaginable health state).

### Measures of potential mediators

Measures of potential mediators consisting of knowledge, goals, behaivoural cuing, intentions and beliefs about consequences adapted from the intervention logic model will be assessed. A mix of seven bespoke and standardised questions will be used: 1)You set goals and keep track of progress; 2) you are motivated to make changes in your daily activities to change your mood; 3) you are willing to make changes in your life to improve your mood; 4) you find it easy to make a plan [to achieve your goals]; 5) you have trouble making plans to help you reach your goals 6) your behaviour influences your future emotions; and 7) If you make positive changes now you will feel better in the future. Each statement will be scored on a 5-point Likert type scale, with responses ranging from completely agree to completely disagree. In addition, participant’s activation will be assessed at 6 and 12 months follow-up with the Premium Abbreviated Activation scale (PAAS) [46]. PAAS is a 5-item scale, adapted from the behavioural activation for depression scale (BADS-SF) [47]. PAAS encompasses Behavioral Activation of five indicators on a scale of 0 (not at all) to 5 (yes, completely) with scores ranging from 0 to 25.

### Healthcare resource use and Economic outcomes

Healthcare resource use will be collected using a modified Client Service Receipt Inventory (CSRI) [48]. Data will be collected on the use of clinic and outpatient facilities, inpatient stays, tests, imaging and medication, with details of specialities Information on other economic outcomes including employment status, productivity loss, catastrophic health spending, household expenditure and household assets will also be collected.

### Statistical analysis

Baseline characteristics will be summarised by the study arm, using means and standard deviations for continuous data and counts and percentages for categorical data. The flow of study participants will be presented in a CONSORT diagram [49] and fidelity with the BA intervention in terms of the number of sessions and duration of the intervention will be detailed. All primary and secondary outcomes will be summarised descriptively at all available time points, including the extent of missing data.

The primary analysis will be on intention to treat (ITT) basis using a linear mixed effects model, comparing PHQ-9 scores between randomised treatment groups at 6 months from a treatment group by time interaction term, adjusting for country and PHQ-9 at baseline as fixed effects and including a random effect for therapists, nested within the treatment group. The model will incorporate outcomes from all available time points. The treatment group difference at 6 months will be presented as a mean estimate with a 95% confidence interval and associated p-value; the difference at 12 months will be reported as a secondary endpoint. Assumptions of the analysis model will be evaluated (normality and data missing at random), and transformation and multiple imputation considered if appropriate. A Complier Average Causal Effects (CACE) analysis will be conducted as a sensitivity analysis, adjusting for compliance with the BA intervention. Estimates of group differences by country will be explored by including a country by treatment group by time interaction term in the above model as a secondary analysis of the primary outcome.

Secondary outcomes will be analysed similarly to the primary analysis model, using linear or logistic mixed models for continuous and binary outcomes respectively. Adverse events will be summarised by treatment arm in terms of the total number of events, adverse events by type and the average number of events per participant. There will be no statistical comparison of these. Full analysis details will be given in a separate pre-specified statistical analysis plan (SAP).

### Process evaluation

A mixed methods process evaluation will aid in understanding the causal mechanisms underpinning the intervention, along with understanding how such interventions might be implemented and scaled up in practice in primary or secondary healthcare services.

In line with recent guidance, we will focus on understanding:

1. Implementation and fidelity of the intervention
2. Mechanisms of impact of the intervention
3. How context affects delivery and outcomes

We will use data collected from other parts of the programme as well as specifically for the process evaluation.

#### Qualitative data collection using a range of methods

We will collect qualitative data from participants, carers and healthcare staff to explore the responses to the intervention, contextual factors and the mechanisms of action. We will use a range of methods in order to explore these factors in depth, to facilitate discussion and triangulate findings. These may include interviews, focus group discussions and participatory methods such as photovoice. In each country, we will collect data from up to 20 participants, 10 carers, 5 facilitators/supervisors and 3 healthcare managerial staff (exact numbers will be decided closer to the time and will also depend on reaching data saturation). We will also collect data from participants and carers in the control arm. Data will be analysed thematically, comparing data from the controls and any overlap in changes and contextual factors.

#### Reports and observations

The CRFs and session audio recordings will capture important information about the dose and fidelity of the intervention, and behaviour change techniques (BCTs) that were implemented.

#### Trial data

Collected at baseline, after completion of BA sessions and at 6- and 12-months post-randomisation looking at potential mediators derived from the intervention logic model will be analysed, to see which are important and changes that may occur during the intervention.

Findings from the process evaluation will be mapped to an implementation framework to explain how the intervention was delivered, the mechanisms of action and contextual factors. Additionally, it will provide opportunities to generate hypotheses that might explain differential responses among groups of participants. This will inform secondary and pre-planned subgroup analysis of the main trial data to further investigate differential treatment responses.

### Economic analyses

Information on the costs and health effects of alternative interventions are needed to inform the allocation of limited healthcare resources to meet population health needs. The economic analysis will provide evidence to consider the value for money of the DiaDeM intervention. Further, it will examine the economic consequences of diabetes and depression more generally.

#### Cost-effectiveness analysis

Cost-effectiveness analysis will examine the value for money of the DiaDeM intervention for individuals with diabetes and depression multimorbidity over the trial period and over the individuals’ lifetime. The within-trial analysis will examine the value for money of the DiaDeM intervention compared to optimised usual care over the 12-month trial period. This will be assessed from a healthcare perspective in the base case. Alternative perspectives reflecting broader economic outcomes will also be considered as scenario analyses [50]. Outcomes will be measured in both quality-adjusted life-years (QALYs) and disability-adjusted life-years (DALYs), generic measures of health which capture both morbidity and mortality[51]. Resource use will be captured, and costs estimated by applying unit costs to resource use. Cost-effectiveness will be presented using incremental cost-effectiveness ratios and incremental net health benefits and net monetary benefits based on appropriate cost-effectiveness thresholds [52,53].

Analyses will control for any baseline differences in covariates between patient groups and for issues with non-normality of cost and outcome data using appropriate statistical techniques [54]. Missing data will be imputed using imputation with chained equations [55].

#### Extrapolation of results over a lifetime

Treatment effects observed in the trial on depression and other risk factors (e.g., HbA1c) will be incorporated into the previously developed DiaDeM depression and diabetes multimorbidity model to estimate the lifetime cost-effectiveness of DiaDeM versus optimised usual care. Evidence on resource use and costs from the trial will also be incorporated into the model.

Cost-effectiveness will be presented using incremental cost-effectiveness ratios and incremental net health benefits and net monetary benefits based on appropriate cost-effectiveness thresholds [52,53]. Scenario analyses will also be conducted.

#### Evaluating economic impacts of depression and diabetes

Using evidence on depression and diabetes status and economic outcomes (including employment status, productivity loss, catastrophic health spending, household expenditure and household assets) collected in the trial, the causal impact of disease on economic outcomes will be assessed. A two-stage modelling approach will be employed. First, we will evaluate the distributional impact of DiaDeM using difference-in-differences (DID) analysis to compare outcomes for the DiaDeM treatment arm with the control arm. To account for heterogeneity in the impact of DiaDeM, we will interact the DID estimator with the wealth level of the household, measured by the asset index. This first stage will allow us to obtain the predicted measure of depression-diabetes multimorbidity. In the second part, to evaluate the causal impact on economic outcomes, we will regress the predicted measure of depression-diabetes multimorbidity from the first stage, on the economic outcome variables. For each outcome, we will run appropriate regressions. We will take account of socio-demographic and other control variables including gender and will run sensitivity checks to ensure the robustness of our models. This approach will help us with the identification problem arising from the fact that several of our economic outcomes can also determine depression (i.e. reverse causality). We will condition on baseline depression status and look at attributed changes in the outcome variable to the intervention, i.e., the mental health of participants which is mediated by the impact of the DiaDeM intervention on their depression.

### Adverse events monitoring

Although we expect adverse or serious adverse events encountered during the trial will be minimal, a standard approach to the collection, recording and reporting of adverse and serious adverse events will be followed in order to ensure the safety of trial participants.

#### Definition of adverse events

Adverse events (AEs) are defined as an untoward clinical development that may occur in an individual during their participation in a clinical trial. This could be an unfavourable and unintended sign, symptom, or disease that may or may not be related to the treatment provided [56]. In our study population of people with diabetes and depression, a range of potential adverse events could be expected. These include, but may not be limited to: hospitalisation for any given cause, diabetes emergencies like the development of cataracts, partial or complete blindness, development of ulcers, hypersensitivity reactions, hypoglycemia, severe hypoglycemia (hypoglycemic shock), diabetic ketoacidosis (DKA) and hyperosmolar hyperglycemic state (HHS) deterioration of renal function leading to chronic kidney disease (CKD), kidney failure, diabetes complications like angina, myocardial infarction, stroke, transient ischemic attacks, loss of sensation in the feet, diabetic foot, gangrene, limb, foot or toe amputations. Depression-related emergencies (suicidal ideation) captured on the PhQ-9 assessment either at baseline or follow-up assessments will also be recorded as an adverse event.

Adverse events will not include medical or surgical procedures other than those identified in the list of potential adverse events above. They will not include pre-existing diseases or hospitalisation in cases where no untoward or unintended response has occurred (elective cosmetic surgery, or social admissions).

#### Identification and reporting of adverse events

Information on adverse events will be collected by RAs as reported by the medical officers at the study sites and/or by the trial participants’ self-report through the course of their involvement in the trial. Information on the type of expected adverse events, along with information on how to report adverse events in a quarterly newsletter will be mailed to all study participants from the month of their recruitment till the end of follow-up activities. Trial participants who contact the study team to report adverse events will be put in touch with the study RA, who will use a pre-designed data collection form to collect the relevant information.

The collected adverse events information will be assessed by a designated member of clinical staff at each study site for expectedness, relatedness, and seriousness. The WHO Uppsala Monitoring Centre (WHO-UMC) scale [57] will be used to identify the relatedness of each unexpected event, which will classify events as certain, probable, possible and unlikely. Seriousness will be assessed on the basis of whether the observed event i) results in death ii) is life-threatening, ii) requires inpatient hospitalisation or prolongation of existing hospitalisation or iv) results in persistent or significant disability or incapacity.

For expected adverse events, a standard reporting procedure will be followed, whereby the information will be reported at scheduled meetings of the Data Monitoring Committee. Adverse events that are classified as serious, unexpected and related to the intervention will require expedited reporting to the Trial Manager and the Principal Investigator who will review the report and inform the York ethics committee within 15 days of the occurrence. Participants reporting such events will be followed up until the episode has resolved or an outcome has been reached. Follow-up information, where applicable, will also be sent as soon as it is available. In addition, each study site staff will follow their institution’s procedure for local notification and as per the guidelines of their local Research Ethics Committees Boards.

### Trial monitoring and quality assurance

We will ensure that our research activities align with internationally-agreed ethical standards [58,59], and the Medical Research Council’s (MRC) guidance on the management of global health trials [60] while at the same time complying with local legislation governing research participation and staff employment in Bangladesh and Pakistan. Research staff will be trained (and re-trained in due course, where necessary) on Good Clinical Practices (GCP), and on key trial procedures such as taking informed consent, administering questionnaires, recording participant responses, and maintaining study logs, ethical considerations, and data management. We will follow the University of York and its partners’ policies for the safety of staff and research participants. Ethics applications and protocols will detail how this will be monitored and risks mitigated. For UK staff travelling to partner countries, risk assessments will be carried out according to university policy. For LMIC staff working in field settings, risks will be assessed by the site lead and any mitigations required will be put in place. We will work within York’s Safeguarding Policy, and in line with the statement on standards for safeguarding international development research.[61]

A Data Monitoring Committee (DMC) comprising independent members will act as the oversight body for the DiaDeM trial. The DMC will safeguard the interests of trial participants, assess the safety of the interventions during the trial, and monitor the overall conduct of the trial. The DMC will decide the frequency of their meetings before and during the trial to review the progress, and quality of data and monitor AEs. They will monitor the safety of study participants and that they are not exposed to unnecessary risks because of their trial participation.

The overall conduct of the trial and its procedures will be monitored regularly by an Independent Trial Steering Committee (ITSC). The ITSC will provide oversight of the trial procedures to ensure the validity and credibility of the trial, and protocol adherence. The ITSC will also be advised on the conduct of the trial with special reference to the safety of the participants and researchers by the DMC after continuously appraising and evaluating the validity and integrity of the trial data as it is accrued. The trial may be stopped if guided by ITSC and DMC, as per termination guidelines.

### Community Engagement and Involvement

Community Advisory panels (CAP) will be convened in each country every 6 months throughout the duration of the trial. Each panel will help to ensure the programme remains focused on the prioritised needs of the community and that planned activities are ethically sound, culturally appropriate, feasible and have community support. Panel members will review plans and study materials for recruitment and data collection; they will also advise on and contribute to the dissemination of key findings to participants, healthcare providers, and public. CAP involvement will improve research quality, help engender a sense of ownership of outputs by the community, help to accelerate uptake and maximise the reach and impact of our programme.

### Ethical considerations

Formal ethical approvals have been acquired from the Health Sciences Research Governance Committee (HSRGC), University of York (Ref: HSRGC/2020/409/B), Diabetic Association of Bangladesh (Ref: BADAS-ERC/EC/20/00300), National Bioethics Committee Pakistan (Ref: No.4-87/NBC-578/23/1382), Institutional Research and Ethics Forum of Rawalpindi Medical University (Ref:242/IREF/RMU/2020) and Ethics Committee of the Office of Research Innovation & Commercialization (ORIC), Khyber Medical University (KMU),(Ref: DIR/KMU/UEC/25), Pakistan.

The study protocol adheres to the Declaration of Helsinki’s principles of human rights and dignity [62]. All research activities will be conducted in accordance with current MRC Good Clinical Practice guidelines [63], the NHS Research Governance Framework [64], and the Mental Capacity Act (UK) 2005 [65]. The latter identifies good practices governing the participation of people lacking capacity. All individuals screened for the trial will be provided an information leaflet, highlighting the importance of depression screening and possible care pathways for those who have depressive symptoms. For those taking part, we will follow standard procedures for informed consent, maintain participant anonymity and explain the right to withdraw at any time during the study. For those who choose to withdraw, we will retain and utilise data gathered up until the day of withdrawal, unless otherwise specified. We will also seek permission to retain contact information for possible contact in the future for other research projects.

Adverse events that need expedited reporting will be notified according to the requirements and within the time periods specified in the Adverse events procedures. In the case of abnormal blood reports, or the detection of a comorbid illness based on biochemical assessment or screening tests/scales, referrals will be made to the relevant medical department. The suicide risk pathway and standard operational procedure developed for each study site will be followed for those who are identified as being at risk of self-harm (Supplementary file 9). All participants will be appropriately reimbursed for their travel for each contact session.

### Data Management

During the trial, quantitative and qualitative data will be collected through researcher-administered interviews, face-to-face assessments, interviews, observations, photos, and focus groups. For screening and blood tests, data will be entered on paper and will then be entered into a secure online, password-protected data entry software. Baseline and follow-up CRFs and information on anthropometric measurements will be entered directly into Qualtrics. This will be done by RAs using handheld tablets at each site. In addition to the above, some DiaDeM intervention sessions will be audio recorded, with consent, for the process evaluation and fidelity assessment of intervention delivery. Interviews and focus group discussions will also be audio-recorded. Written notes will also be recorded in hard copies along with screening forms, consent forms and blood reports. Participant logs for completion of fidelity indices, follow-up and withdrawal will be recorded using standard electronic templates. All collected data will be stored on a central server at the University of York.

Qualitative interviews will be transcribed in the interview language, but linked with the trial ID and will be anonymised before translation for analysis. Digital recordings of the interviews will be stored locally on a temporary basis in a password-protected folder. Once the analysis of the interviews is completed all recorded versions will be erased from digital recorders.

The Trial Statistician/data manager based at the University of York and the country trial data managers will have access to the datasets. They will use verification, validation, and checking procedures to ensure that the data are accurate and comprehensive. All members of the research team will adhere to the data management policy developed for DiaDeM under the direction of the University of York Trials Unit’s Standard Operating Procedures. The research team will ensure the confidentiality and anonymity of participants to those with delegated responsibilities within the trial team. Participants will be assigned a unique ID so that no identifiable information is accessible to non-authorized persons. Moreover, all identifiable information of the participants will be stored at each study site securely; electronic records will be kept under password protection and hard copies will be stored in locked filing cabinets.

Secure services available through the University of York (University of York drop-off: https://www.york.ac.uk/it-services/services/dropoff/) will be used to transfer encrypted data between study sites and the University of York, where necessary. All files transported through this will be encrypted as zip files using passwords consisting of at least 8 characters with a mix of letters and numbers in upper and lower case, and punctuation or other symbols. Data received via drop-off will only be accessed on desktop computers or laptops which are connected to an encrypted server. Moreover, for non-confidential data or non-sensitive data, researchers will share the data through the DiaDeM team drive on the Google Drive cloud-based storage platform, or through the University of York <u>drop-off</u> service.

In line with the University of York Research Data Management Policy, all anonymised research data that underpin published results or have long-term value will be retained indefinitely after the completion of trial activities. Upon completion of the DiaDeM activities, all non-anonymised data will be destroyed.

## Discussion

The DiaDeM trial aims to test the potential of a culturally adapted behavioural activation intervention (DiaDeM) as a suitable and cost-effective treatment to address depression among individuals with Type 2 Diabetes who are living in South Asian settings. We have identified an evidence-based intervention and further adapted it for cultural appropriateness in South Asian users and settings [28]. Our plans for implementing the trial are based on a robust study design, backed by the use of validated measures for outcome evaluation. Findings of our stand-alone feasibility trial of DiaDeM [14] are yet to be published, however, the key message from this work is that the DiaDeM intervention is feasible to deliver in both Bangladesh and Pakistan and is acceptable to both users (to receive) and BA facilitators (to deliver). We have built on the experiences and learning from the feasibility trial by further strengthening our training and supervision processes for BA facilitators. We have further refined the intervention materials (eg. the use of more appropriate images/photographs) based on feedback from research staff and participants. No changes were proposed for the intervention content. We have also expanded the trial to a third location (BIDE, Karachi), which will ensure a more representative sample in Pakistan.

The DiaDeM trial is the first of its kind to be implemented in South Asia. It uses a task-sharing approach, incorporating non-mental health resources available within existing health facilities to address the growing issue of physical and mental multimorbidity. This approach has great relevance for South Asia given its potential for scalability as a feasible and potentially cost-effective solution. We anticipate that our real-world evaluation of the DiaDeM intervention will fill in the existing gaps, paving the way for DiaDeM’s adaptation, implementation, and further evaluation beyond South Asian settings, with a view to improving health-related and economic outcomes in low-resource settings and populations.

## Data Availability

No datasets were generated or analysed during the current study. All relevant data from this study will be made available upon study completion.

## Acknowledgements

All authors were involved in the development of this protocol, and therefore the author sequence is based on an equal contribution approach [66]. We would also like to acknowledge the DiaDeM Program Steering Committee and the Data Monitoring Committee for their oversight, and the Community Advisory Panels in Bangladesh and Pakistan for their input in the development of the trial protocol.

## Funding

This study is being carried out under the National Institute of Health Research, Global Health Research project, [Grant reference: Research & Innovation for Global Health Transformation (RIGHT) NIHR200806]. The University of York is the trial sponsor. The views expressed in this publication are those of the author(s) and not necessarily those of the NIHR or the UK Department of Health and Social Care. The funders had no role in study design, data collection and analysis, decision to publish, or preparation of the manuscript.

## Conflicts of Interest

Authors do not have any conflicts of interests to declare.

## List of supplementary files

S1-SPIRIT checklist

S2-TIDieR checklist

S3-Information leaflet on the recognition of depression

S4-Consent form for stage 2 screening

S5-Participant information sheet

S6-Consent form for trial participation

S7-DiaDeM logic model

S8-Optimised usual care leaflet

S9-Suicide risk pathway and referral procedure

## Notes

### Competing Interest Statement

The authors have declared no competing interest.

### Clinical Trial

ISRCTN40885204

### Author Declarations

Formal ethical approvals have been acquired from the Health Sciences Research Governance Committee (HSRGC), University of York (Ref: HSRGC/2020/409/B), Diabetic Association of Bangladesh (Ref: BADAS-ERC/EC/20/00300), National Bioethics Committee Pakistan (Ref: No.4-87/NBC-578/20/ 1101), Institutional Research and Ethics Forum of Rawalpindi Medical University (Ref:242/IREF/RMU/2020) and Ethics Committee of the Office of Research Innovation & Commercialization (ORIC), Khyber Medical University (KMU),(Ref: DIR/KMU/UEC/25), Pakistan.

